# Subgroups of people with high Autism-Spectrum Quotient scores identified from a large set of personality traits and attributes

**DOI:** 10.1101/2022.07.06.22277323

**Authors:** Mao Fujiwara, Shotaro Numano, Toshiko Tanaka, Motoaki Nakamura, Nobumasa Kato, Masahiko Haruno

## Abstract

The Autism-Spectrum Quotient (AQ) is a widely used index to quantify autistic traits. Previous studies using clustering analysis have indicated that people showing high AQ and other autism indices may comprise several subtypes. However, those studies mainly focused on Big5 personality traits and analyzed a limited number of samples (participants). The inclusion of a larger set of personality traits and attributes would contribute not only to understanding autism more deeply, but also to characterizing AQ more precisely. In addition, an analysis of larger general populations would give new perspectives on people with high AQ scores. To address these issues, here we conducted a personality traits-based clustering of 1364 subjects who had an AQ score of 33 or higher (high AQ group) collected online. We identified three subclasses (subtypes): Group 1, characterized by emotional instability, low self-esteem, high hostility, anxiety, depressive tendency and schizotypal traits; Group 2, characterized by high BIS for an inhibitory attitude towards novelty, and high Interpersonal Reactivity Index (IRI) and Group 3, characterized by personality traits and attributes consistent of the average of the general population. Thus, this study provides empirical results showing subtypes of the high AQ population.

## Introduction

Autism spectrum disorder (ASD) is a developmental disorder characterized by deficits in social communication and repetitive behaviors, including Asperger syndrome and pervasive developmental disorder, in addition to typical infantile autism. In general, ASD is seen in about 1% of the population and is 3∼4 times more common in males than females [1]. The characteristics of ASD are on a continuum from clinical patients to normal individuals [2–4].

The Autism-Spectrum Quotient (AQ) was developed as a self-report questionnaire measure to quantify autistic characteristics [5]. Generally, the AQ is higher in males than in females and in scientists than in non-scientists [5, 6]. Adults with Asperger’s syndrome and high-functioning autism (AS/HFA) score significantly higher on the AQ than adults in the general population. These observations indicate that the AQ is a valid measure of autistic traits [5, 7]. As for the relationship between the AQ and other disorder-related personality traits, there are relatively strong positive correlations between measures of social skills and communication in the AQ and the four sub-items of the Schizotypal Personality Questionnaire (SPQ)-interpersonal factors (social anxiety, suspicion, friendlessness, and emotional suppression) [8]. In addition, almost half of women with borderline personality disorder (BPD) show high AQ scores, and comorbid BPD correlates with having a higher AQ score [9].

The relationship between the AQ and Big5, which describes basic personality traits, has been long studied. Wakabayashi et al. [7] found that AQ scores are negatively correlated with extraversion and conscientiousness, positively correlated with neuroticism, and not correlated with agreeableness. In contrast, Austin [6] reported that the AQ was negatively correlated with agreeableness. A recent meta-analysis of Big5 in ASD not only validated a negative association between each Big5 trait and ASD characteristics, but it also emphasized the importance of extending the relationship of ASD with personalities beyond Big5 such as attachment style, emotion regulation, alexithymia, and self-esteem [10].

Related to the above observations, there are many cases of the co-occurrence of autism with anxiety disorders (43-84%), depression (2-30%), obsessive-compulsive disorder (OCD; 37%), attention-deficit/hyperactivity disorder (59%), agitation and aggressive behavior (8-34%), and nonspecific abnormal behaviors, such as self-injury [11]. The data of these broad comorbidities and variable co-occurrence rates suggest that ASD may consist of several subclasses, and questionnaires about ASD need to link with these disorders. Such classes, if any, would contribute not only to understanding ASD more deeply and developing future intervention methods for each class, but also to characterizing ASD indices such as AQ more precisely.

Indeed, to identify the subclasses, several studies have conducted a clustering of people with high ASD scores, although the majority of studies have treated this population with a high ASD score as a single group [12, 13]. A study using the RAADS-R score (an index of ASD) identified four subtypes based on the Big5 factors. Specifically, cluster 1 has Big5 features similar to social phobia [11], and cluster 2 shows a Big5 score distribution similar to secondary psychopath and BPD [12]. Cluster 2 and cluster 3 have low agreeableness, extraversion, and average levels of emotional stability and conscientiousness but high happiness. Cluster 4 has the highest extraversion, agreeableness, and happiness among the four clusters [13]. That study examined 364 individuals with high ASD traits recruited from autism-related websites and based on the results of Big5 factors and subcategories, life satisfaction, and happiness. Another study clustered 2000 autistic individuals based on 123 item scores from the Autism Diagnostic Interview-Revised (ADI-R) [14] and reported four phenotypic clusters: the first associated with severe language deficits, the second with milder symptoms across the domains, the third with a higher frequency of savant skills, and the fourth with intermediate severity across all domains.

However, the clustering of these previous studies was based on the Big5 or an ASD index alone. In other words, the relationship between the AQ and personality traits other than the Big5 remains largely unknown. In addition, most studies were based on young populations, such as children and undergraduate students; the limited age range makes it difficult to draw general conclusions [6, 15]. Furthermore, when examining general personality types, small sample sizes and the lack or reproducibility across data sets and methods have led to inconclusive results [16]. These findings suggest that a clustering analysis of a larger set of personality traits and attributes beyond the Big5 obtained from a larger and broader population offers useful insights about subclasses in the population with a high ASD score.

Here, we examined online data on AQ and a wide range of personality traits and attributes, including the Big5, psychopathy, schizophrenia, depression, OCD, and other mental illness indicators from a more general population (n=18948), and conducted a co-clustering analysis of 1364 individuals who had an AQ score of 33 or higher (high AQ group) to identify subgroups within this population. To guarantee the generality of the results, we used two distance measures for clustering with the Ward method: the Euclidean distance and the Manhattan distance.

## Results

### The number of clusters in the high AQ group was 3

We first conducted a Lasso regression to select personality traits and attributes that contributed to the AQ score. We then conducted a co-clustering of 1364 subjects with an AQ score ≥ 33 using 35 personality traits (and attributes) whose absolute value of the weights in the Lasso regression was greater than 0.0126, which amounted to 1/20 of the largest absolute value (Table 1) using the Ward method with two different distance measures: the Euclidian and Manhattan distances. We found the optimal number of clusters for this group was 3 for both measures (Fig 1). We show the results only for the Euclidian distance hereafter, as the clusters were comparable between the two distance measures (see S1 Fig. for the cluster structure from the Manhattan distance).

**Fig 1.**
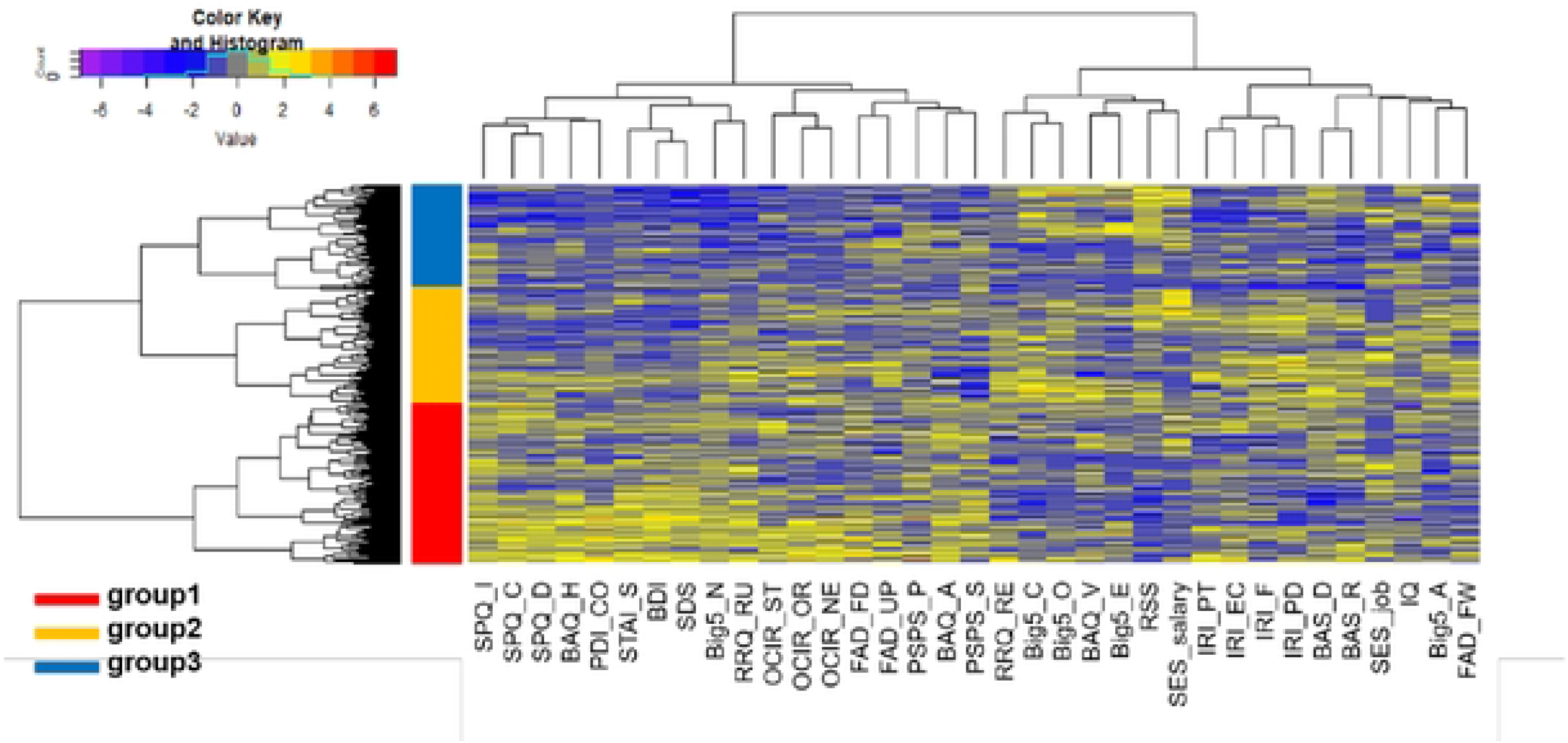
Three clusters in the high AQ group. The horizontal and vertical axes represent the personality traits used for the clustering and the subjects, respectively, with each square showing a standardized score of the corresponding horizontal personality trait of the subject. Color scale: red, high score; purple, low score. For personality traits on the horizontal axis, three blocks were identified: block 1 consisted of BAQ, SPQ, BDI, OCIR, FAD_FD, and UP; block 2 consisted of Big5 and RSS; and block 3 consisted of BIS/BAS and IRI. The high AQ group was clustered into three subgroups: 575 people for Group 1, 410 for Group 2, and 379 for Group 3.

**Table 1.** Personality traits and Lasso regression weights used to determine the number of clusters.

| Personality | Coefficient | Personality | Coefficient | Personality | Coefficient |
| --- | --- | --- | --- | --- | --- |
| Big5_E<sup>c, k</sup> | 0.25257 | IQ<sup>c</sup> | 0.03040 | BIS<sup>k</sup> | 0.00810 |
| SPQ_I<sup>c, k</sup> | 0.24391 | IRI_EC<sup>c, k</sup> | 0.02984 | RA | 0.00808 |
| SPQ_C<sup>c, k</sup> | 0.20272 | RSS<sup>c, k</sup> | 0.02859 | BAQ_P<sup>k</sup> | 0.00497 |
| SPQ_D<sup>c, k</sup> | 0.12839 | BAS_D<sup>c, k</sup> | 0.02794 | SES_parents | 0.00378 |
| Big5_O<sup>c, k</sup> | 0.11126 | PDI_CO<sup>c</sup> | 0.02745 | TIM | 0.00269 |
| Big5_A<sup>c, k</sup> | 0.09556 | SES_job<sup>c</sup> | 0.02532 | SVO_C | 0.00133 |
| FAD_FW<sup>c</sup> | 0.07100 | BAQ_H<sup>c, k</sup> | 0.02325 | OCIR_WA<sup>k</sup> | 0 |
| SDS<sup>c, k</sup> | 0.06943 | OCIR_ST<sup>c, k</sup> | 0.02022 | OCIR_OB<sup>k</sup> | 0 |
| Big5_N<sup>c, k</sup> | 0.06760 | PSPS_P<sup>c</sup> | 0.01909 | PDI_DE | 0 |
| BAS_R<sup>c, k</sup> | 0.06725 | RRQ_RE<sup>c</sup> | 0.01834 | PDI_DI | 0 |
| BAQ_A<sup>c, k</sup> | 0.06720 | PSPS_S<sup>c</sup> | 0.01755 | PDI_FR | 0 |
| OCIR_OR<sup>c, k</sup> | 0.06683 | STAI_S<sup>c, k</sup> | 0.01557 | PSS | 0 |
| IRI_PD<sup>c, k</sup> | 0.06514 | FAD_FD<sup>c</sup> | 0.01546 | SES_self | 0 |
| OCIR_NE<sup>c, k</sup> | 0.06329 | FAD_UP<sup>c</sup> | 0.01521 | SES_subj | 0 |
| BDI<sup>c, k</sup> | 0.05434 | BAQ_V<sup>c, k</sup> | 0.01371 | SHS<sup>k</sup> | 0 |
| IRI_F<sup>c, k</sup> | 0.05109 | SES_salary<sup>c</sup> | 0.01269 | SVO_P | 0 |
| Big5_C<sup>c, k</sup> | 0.04368 | OCIR_CH<sup>k</sup> | 0.01257 | SVO_I | 0 |
| RRQ_RU<sup>c</sup> | 0.03574 | FAD_SD | 0.01169 | STAI_T<sup>k</sup> | 0 |
| IRI_PT<sup>c, k</sup> | 0.03423 | BAS_S<sup>k</sup> | 0.00911 |  |  |

Absolute values of the coefficient (0.0126) amounted to 1/20 the largest absolute value of weights in the Lasso regression analysis (in this case, the largest value was assigned to Big5\_E). The personality traits with values larger than 0.0126 were used both for the clustering and for determining the number of clusters.
<sup>c</sup> what we used for clustering, <sup>k</sup> what we used for kernel density estimation

The three groups are summarized as follows. In comparison with the mean values of the groups, Group 1 showed slightly higher scores on OCIR [17], BDI [18], SPQ [19], BIS [20], IRI [21] and STAI [22] and lower scores on the Big5 [23] other than Big5_N. Group 2 showed higher scores on BIS, the Big5 and IRI, and slightly higher scores on personality traits related to mental disorders, such as OCIR, SPQ. Lastly, Group 3 exhibited lower scores on OCIR, BDI and SPQ.

### Characteristics of the three high AQ subgroups

An analysis of variance revealed that 52 out of 61 personality trait (sub-) scores including the AQ showed significant differences among the three groups (S2 Table). Figs 2-7 plot the probability density distribution of the personality trait scores for each group. In these figures, the vertical and horizontal axes show the percentage of the number of people and the scores, respectively, and p-values were calculated using Welch’s T-test. The threshold for significance was set at 0.0002 (0.05 divided by the number of items used for the clustering, 35, and the combination of groups, 6). Considering that the p-value tends to decrease with a larger sample size, this threshold was considered a minimum requirement.

**Fig 2.**
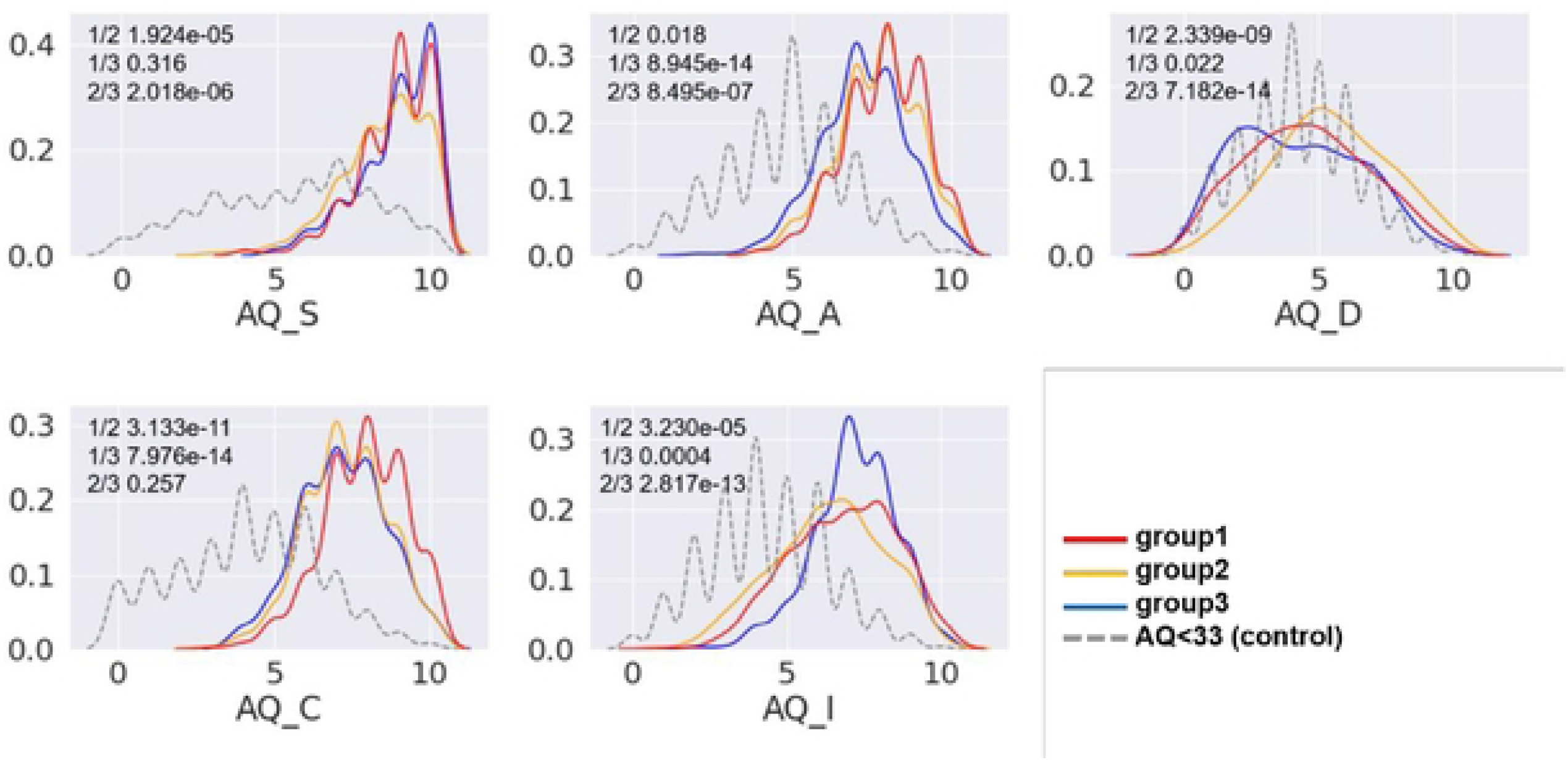
AQ scores for each group. For AQ_S, Group 2 had a significantly higher mean value than Groups 1 and 3 (Group 2 vs. 1, f=4.300, p=1.924e-05; Group 2 vs. 3, f=-4.787, p=2.018e-06). For AQ_A, Group 3 had a significantly lower mean value than Groups 1 and 2 (Group 3 vs. 1, f=7.594, p=8.945e-14; Group 3 vs. 2, f=4.964, p=8.945e-07). For AQ_D, Group 2 had a significantly higher mean value than Groups 1 and 3 (Group 2 vs. 1, f=-6.033, p=2.339e-09; Group 2 vs. 3, f=7.624, p=7.182e-14). For AQ_C, Group 1 had a significantly higher mean value than Groups 2 and 3 (Group 1 vs. 2, f=6.726, p=3.133e-11; Group 1 vs. 3, f=7.608, p=7.976e-14). For AQ_I, Group 3 had a significantly higher mean than Groups 1 and 2 (Group 3 vs. 1, f=-3.574, p=0.0004; Group 3 vs. 2, f=-7.436, p=2.817e-13). AQ_S: social skills, AQ_A: attention switching, AQ_D: attention to detail, AQ_C: communication, and AQ_I: imagination). Higher scores indicate stronger autistic traits; the higher the score, the stronger the autistic trait. In other words, a higher AQ_S means poorer social skills, a higher AQ_A means poorer attention switching, a higher AQ_D means higher attention to detail, a higher AQ_C means poorer communication, and a higher AQ_I means poorer imagination.

Fig 2 shows the distribution of AQ scores for each group. Group 1, Group 2 and Group3 have higher AQ_S, AQ_A, AQ_I, and AQ_C compared to the general population and overall have high autistic tendencies. Group 3 is characterized by high AQ_I.

Fig 3 shows emotion-related items. BDI and SDS [26] measure depressive tendencies, STAI_S measures anxiety as a transient mood by asking how one feels about oneself at the time of the measurement, and STAI_T measures one’s susceptibility to anxiety by asking how one usually feels about oneself [27]. Group 1 had higher depressive tendency and anxiety than the other groups and the general population, suggesting that members of this group tended to be mentally unstable. In Group 3, both depression and anxiety were much lower than in Group 1, and they tended to be mentally calm. Group 2 was similar to Group 3, but STAI_T was higher than in Group 3. In addition, Group 1 had the lowest self-esteem and the lowest happiness among the three groups. Fig 3 also shows the distribution of BAQ [24] scores, an index of aggression. Group 1 is characterized by high scores on BAQ_P, BAQ_A, and BAQ_H. BAQ_P is a scale measuring physical aggression reactions, with items measuring the tendency to violent reactions, impulses to violence, and justification of violence. BAQ_A is a scale measuring anger arousal, with items measuring anger and low anger inhibition. BAQ_H is a scale measuring suspicion and distrust, including malice and disregard for others, with items measuring negative beliefs and attitudes toward others. BAQ_V is a scale that measures verbal aggression reactions and consists of items that measure assertiveness and argumentativeness [25]. Therefore, Group 1 is highly suspicious and distrustful, short-tempered, and has little resistance to violence. Group 2 is characterized by high scores on BAQ_A and BAQ_H. Like Group 1, this group is short-tempered and paranoid. By contrast, Group 3’s BAQ scores were comparable to the general population.

**Fig 3.**
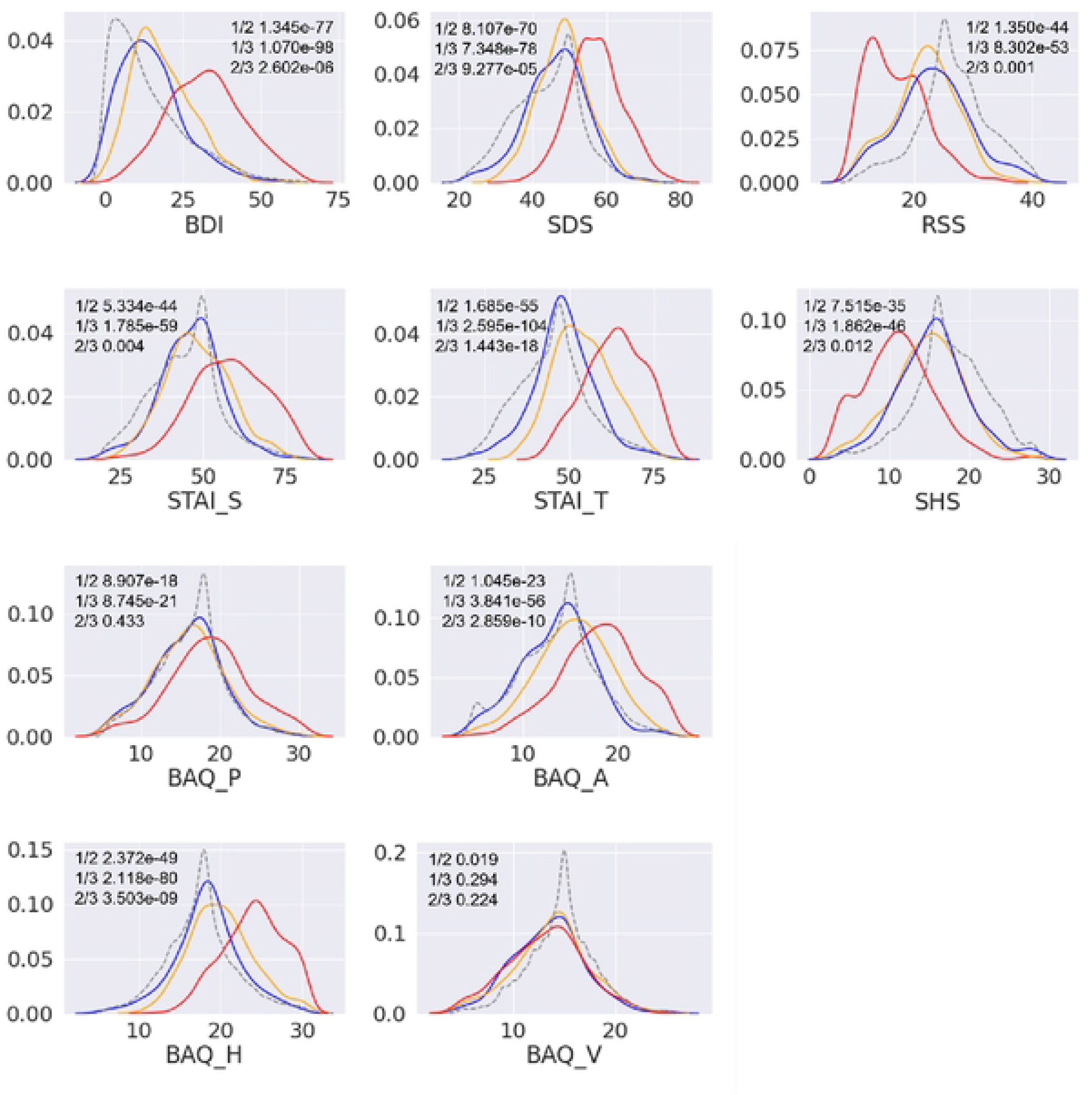
Emotion-related items for each group. For BAQ_P, BDI and SDS, Group 1 had significantly higher mean values than Groups 2 and 3 (BDI: Group 1 vs. 2, f=20.451p=1.345e-77; Group 1 vs. 3 f=24.005, p=1.070e-98; and SDS: Group 1 vs. 2, f=19.305, p=8.107e-70; Group 1 vs. 3, f=21.119, p=7.348e-78). Group 1 also had significantly lower mean values for STAI_S and STAI_T than Groups 2 and 3 (STAI_S: Group 1 vs. 2, f=14.662, p=5.334e-44; Group 1 vs. 3, f=17.547, p=1.785e-59; and STAI_T: Group 1 vs. 2, f=16.849, p=1.685e-55; Group 1 vs. 3, f=25.416, p=2.595e-104). For RSS, Group 1 had significantly lower mean values than groups 2 and 3 (RSS: Group 1 vs. 2, f= −14.869, p= 1.350e-44; Group 1 vs. 3 f= −16.700, p= 8.302e-53). For SHS, Group 1 had significantly lower mean values than groups 2 and 3 (SHS: Group 1 vs. 2, f= −12.871, p= 7.515e-35; Group 1 vs. 3 f= −15.277, p= 1.8612e-46). BDI: depression, SDS: depression, STAI_S: state anxiety, STAI_T: trait anxiety, RSS: self-esteem, and SHS: subjective happiness. For BAQ_A and BAQ_H, Group 1 had significantly higher mean values than Groups 2 and 3 (BAQ_P: Group 1 vs. 2, f=8.762, p=8.907e-18; Group 1 vs. 3, f=9.587, p=8.745e-21; BAQ_A: Group 1 vs. 2, f=10.325, p=1.045e-23; Group 1 vs. 3, f=17.013, p=3.841e-56; and BAQ_H: Group 1 vs. 2, f=15.761, p=2.372e-49; Group 1 vs. 3, f=21.482, p=2.118e-80). For BAQ_V, Group 1 had no significant difference with Groups 2 or 3 (Group 1 vs. 2, f=-2.344, p=0.019; Group 1 vs. 3, f=-1.050, p=0.294). BAQ_P: physical aggression, BAQ_A: short temper, BAQ_H: suspicion, and BAQ_V: verbal aggression.

Fig 4 displays the results for the Big5 and IQ. Big5_E, Big5_A, Big5_C, Big5_N, and Big5_O measure extraversion, agreeableness (concerning compassion and kindness), conscientiousness (concerning perseverance and thoroughness), neuroticism (concerning meticulousness and emotional instability) and openness (concerning the ability to take in a wide range of experiences and make decisions), respectively, and IQ measure reasoning ability. Group 1 is characterized by a higher Big5_N than the other groups, indicating that Group 1 is less emotionally stable. Group 2 is characterized by higher levels of Big5_A, Big5_C, and Big5_O than the other two groups, meaning that Group 2 is as kind and perseverant as the general population. In addition, Group 2 had a slightly higher average IQ. Group 3, like the other two groups, is low in extraversion and shows the general tendency of people with high autistic scores, but it is also characterized by the lowest Big5_N among the high AQ groups and is therefore considered to be the most emotionally stable among the three groups.

**Fig 4.**
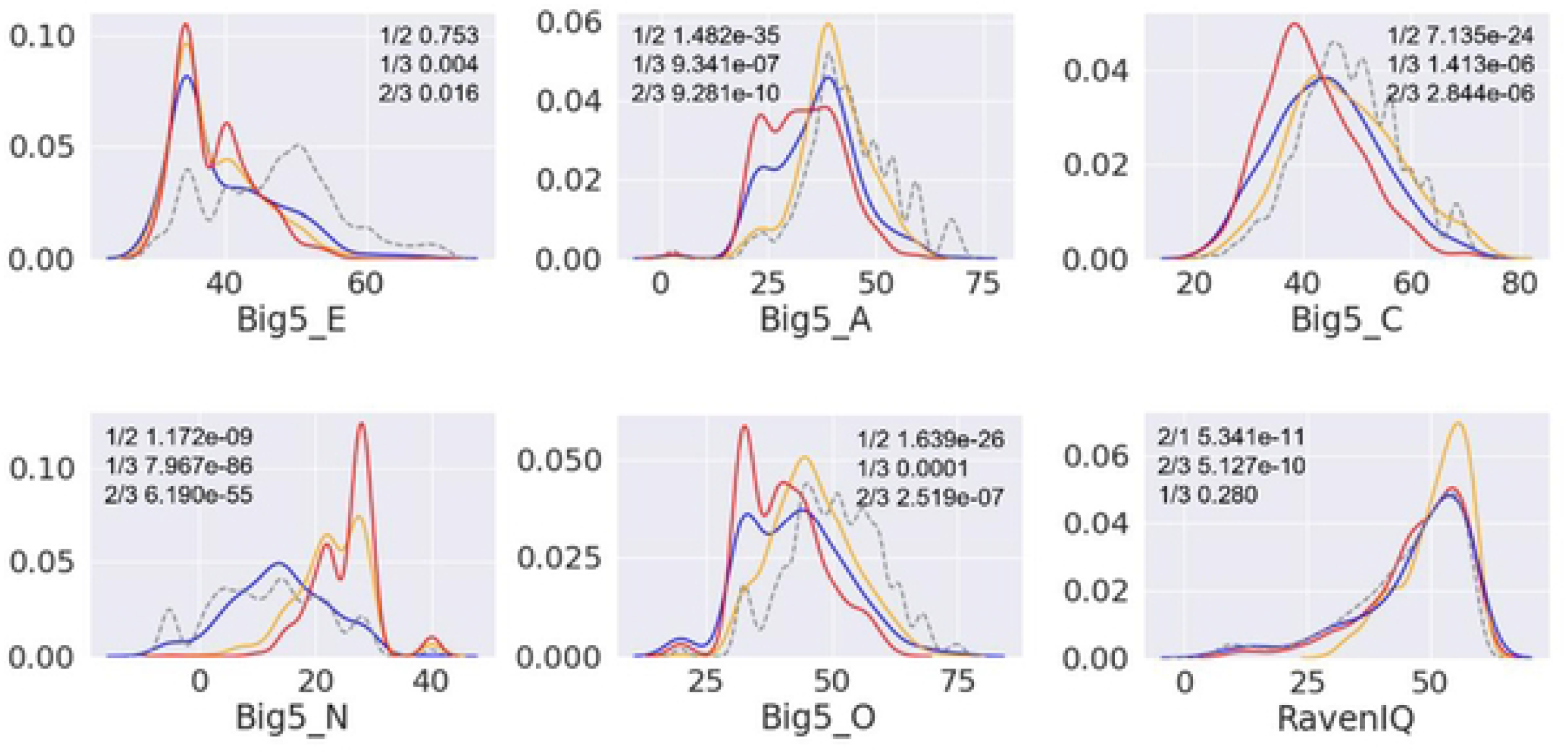
Big5 for each group. For Big5_E, Group 2 had no significant difference with Groups 1 or 3 (Group 2 vs. 1, f=-0.314, p=0.753, Group 2 vs. 3, f=-2.417, p=0.016). For Big5_A, _C, and _O, Group 2 had significantly higher mean values than Groups 1 and 3 (Big5_A: Group 2 vs. 1, f=-13.001, p=1.482e-35; Group 2 vs. 3, f=6.202, p=9.281e-10; Big5_C: Group 2 vs. 1, f=-10.410, p=7.135e-24; Group 2 vs. 3, f=4.716, p=2.844e-06; and Big5_O: Group 2 vs. 1, f=-11.016, p=1.639e-26; Group 2 vs. 3, f=5.205, p=2.519e-07). For Big5_N, Group 3 had a significantly lower mean value than Groups 1 and 2 (Group 3 vs. 1, f=23.443, p=7.967e-86; Group 3 vs. 2, f=17.100, p=6.190e-55). Big5_E: extroversion, Big5_A: cooperativeness, Big5_C: diligence, Big5_N:neuroticism, and Big5_O: openness. For IQ, Group 2 had a significantly higher mean value than Groups 1 and 3 (Group 2 vs 1: f = −6.635, p = 5.341e-11; Group 2 vs 3: f = 6.320, p = 5.127e-10; Group 1 vs 3: f = 1.081, p = 0.280). IQ: intelligence quotient.

Fig 5 shows OCD-related and schizophrenia-related scores. For the OCIR, which measures the frequency of OCD symptoms, Group 1 scored higher than the other groups on all items. SPQ measures the characteristics of schizotypal personality and consists of nine subtypes of schizophrenia: SPQ_C, SPQ_I, and SPQ_D are indices for relational ideation, magical thinking, abnormal perception, and suspicion; social anxiety, absence of friends, emotional suppression, and suspicion; and odd behavior and odd speech, respectively. Example questionnaires for each item were “I have to be careful because people may resent me or have bad intentions toward me, or my friends may actually be dishonest,” “I would rather be alone than interact with others, and I am not good at socializing,” and “I am not good at speaking expressively or expressing my true feelings” [28]. The questions in the survey overlap with those in BAQ_H for suspicion and AQ_S for social anxiety and absence of close friends. All SPQ scores were predominantly higher for Group 1 than the two groups, but the SPQ_I scores were higher than in the general population not only in Group 1 but also Group 2 and Group 3, suggesting that high AQ people have difficulty with interpersonal relationships. In addition, all groups scored high on AQ_S and AQ_C. Group 3 had the lowest scores on all items.

**Fig 5.**
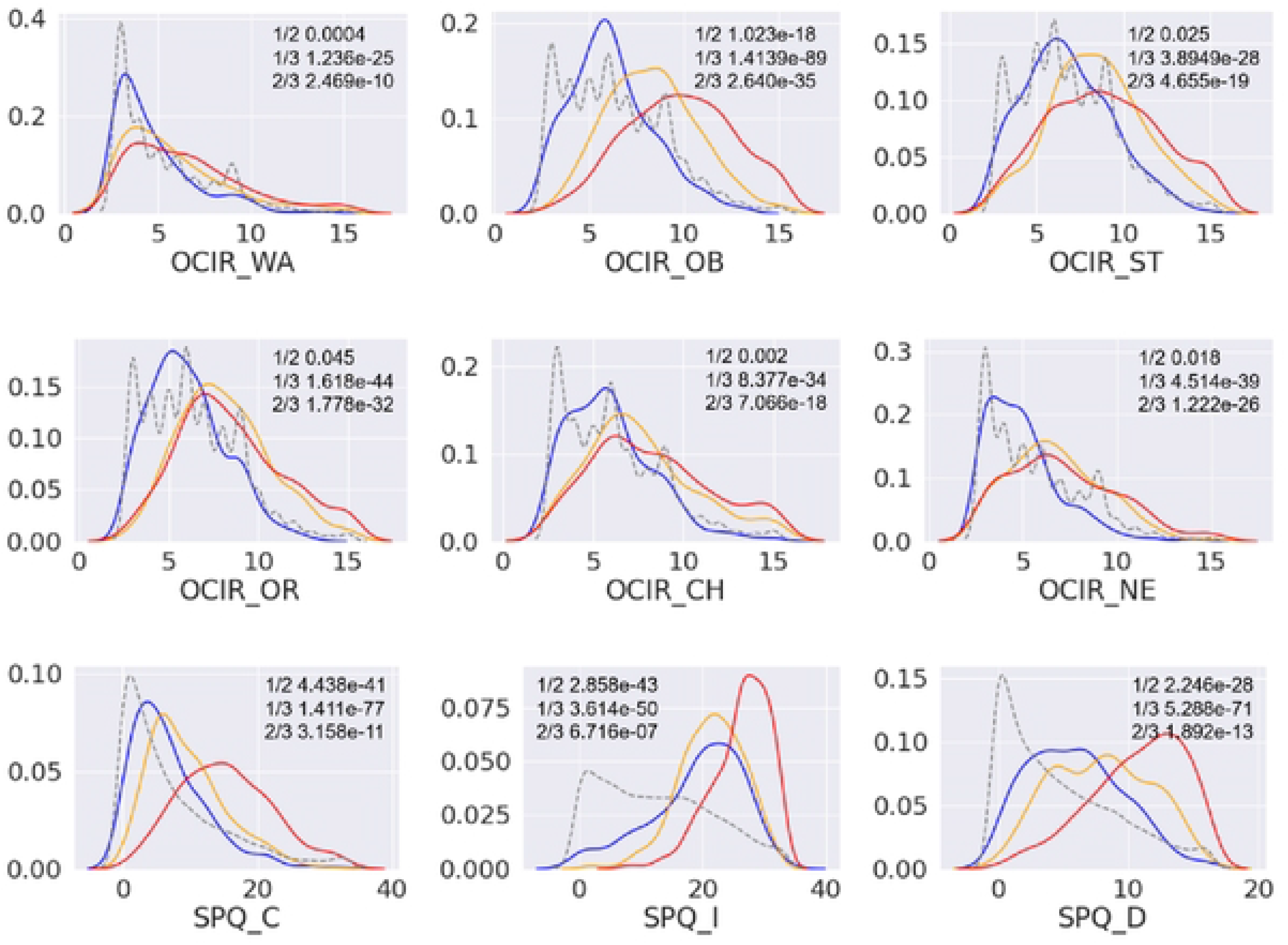
OCD-related scores and schizophrenia-related scores for each group. Group 3 had significantly lower mean values than Groups 1 and 2 for OCIR_WA, OCIR_ST, OCIR_OR, OCIR_CH and OCIR_NE (OCIR_WA: Group 3 vs. 1, f=10.779, p=1.236e-25; Group 3 vs. 2, f=6.416, p=2.469e-10; OCIR_ST: Group 3 vs. 1, f=11.370, p=3.8949e-28; Group 3 vs. 2, f=9.154, p=4.655e-19; OCIR_OR: Group 3 vs. 1, f=14.759, p=1.618e-44; Group 3 vs. 2, f=12.428, p=1.778e-32; OCIR_CH: Group 3 vs. 1, f=12.608, p=8.377e-34; Group 3 vs. 2, f=8.830, p=7.066e-18; and OCIR_NE: Group 3 vs. 1, f=13.690, p=4.514e-39; Group 3 vs. 2,f=11.095, p=1.222e-26). For OCIR_OB, Group 1 had significantly higher mean values than Groups 2 and 3 (Group 1 vs. 2, f=9.019, p=1.023e-18; Group 1 vs. 3, f=22.454, p=1.4139e-89) OCIR_WA: wash, OCIR_OB: obsess, OCIR_ST: store, OCIR_OR: order, OCIR_CH: confirm, and OCIR_NE: number. Group 1 had significantly higher mean values than Groups 2 and 3 for all SPQ items (SPQ_C: Group 1 vs. 2, f=14.080, p=4.438e-41; Group 1 vs. 3, f=20.578, p=1.411e-77; SPQ_I: Group 1 vs. 2, f=14.636, p=2.858e-43; Group 1 vs. 3, f=16.457, p=3.614e-50; and SPQ_D: Group 1 vs. 2, f=11.455, p=2.246e-28; Group 1 vs. 3, f=19.712, p=5.288e-71). SPQ_C: cognitive and perceptual, SPQ_I: interpersonal, and SPQ_D: demolition.

Fig 6 shows the results for BIS and BAS. BIS concerns the avoidance of conditioned aversive stimuli, and BAS activates behavior in response to incentives. BAS has three subscales, BAS_D, BAS_R, and BAS_S, representing a persistent pursuit of goals (e.g., “I will do anything to get what I need”), positive responses to rewards (e.g., “I get excited and energized when I get what I want”), and desire for new rewards (e.g., “I’m eager to get excitement and new sensations”), respectively [29]. Group 2 had a higher BAS_D score than the other two groups. According to McCrae & Costa [30], neuroticism is a personality trait that reflects the ease of experiencing negative emotions, while extraversion is a vivacious personality trait such as being socially active and seeking excitement and stimulation. Takahashi et al. [31] found a positive correlation between BIS and neuroticism and a positive correlation between BAS and extraversion. In the present study, Group 1 had a higher BIS and higher neuroticism tendency (high Big5_N, i.e. low emotional stability), but there was no difference in the distribution of Big5_E scores between the three groups despite the differences in the distribution of BAS scores. Group 2 showed a similar level of BIS to Group 1. Group 3 had the same mean and peak score as the average of all subjects for all items.

**Fig 6.**
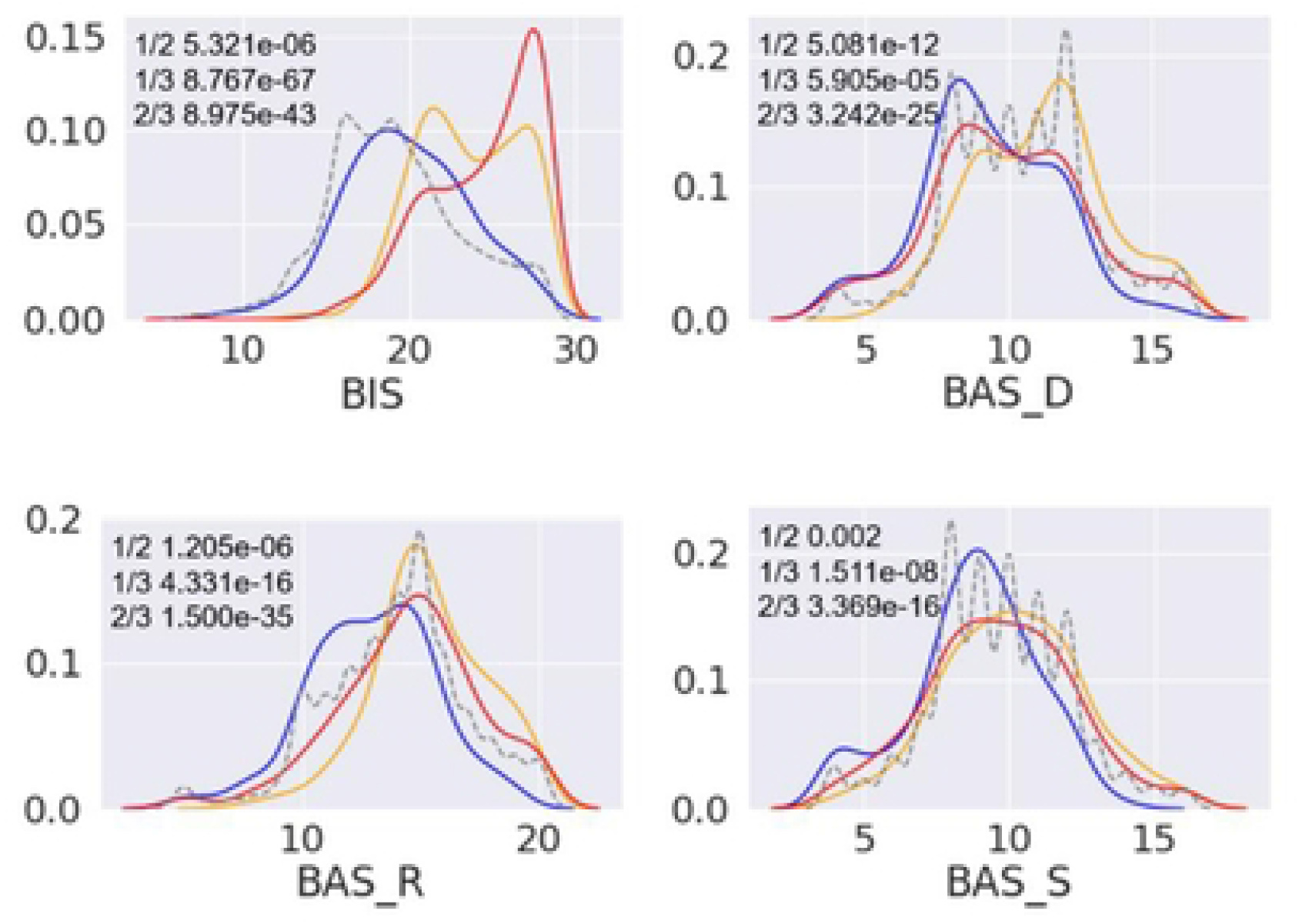
Behavioral inhibition system (tendency to avoid punishment) and behavioral activation system (tendency to approach reward) for each group. For BIS, Group 3 had a significantly lower mean value than Groups 1 and 2 (Group 3 vs. 1, f=19.186, p=8.767e-67; Group 3 vs. 2, f=14.641, p=8.975e-43). For BAS_D, Group 2 had a significantly higher mean value than Groups 1 and 3 (Group 2 vs. 1, f=-6.993, p=5.081e-12; Group 2 vs. 3, f=10.747, p=3.242e-25). For BAS_R and BAS_S, Group 3 had a significantly lower mean than Groups 1 and 2 (BAS_R: Group 3 vs. 1, f=8.288, p=4.331e-16; Group 3 vs. 2, f=13.109, p=1.500e-35; BAS_S: Group 3 vs. 1, f=5.713, p=1.511e-08; Group 3 vs. 2, f=8.338, p=3.369e-16). BIS: behavioral inhibitory system, BAS_D: behavioral activating system_driven, BAS_R: behavioral activating system_reward responsive, and BAS_S: behavioral activating system_stimulus seeking.

Fig 7 shows the results for the IRI, which measures empathy. IRI_EC, IRI_PT, IRI_PD, and IRI_F represent “the ease with which other-oriented emotions such as sympathy are aroused,” “the degree to which one considers the feelings of others from their perspective,” “the degree to which one is preoccupied with anxiety or fear generated in oneself by observing the distress of others,” and “the tendency to imagine oneself as a character in a fictional story,” respectively [32]. For IRI_PT and IRI_EC, Group 1 and 2 scored higher than the general population, and for IRI_F, IRI_PD, Group 2 scored significantly higher than the other two groups. Group 3 showed scores consistent of the general population for all items of IRI.

**Fig 7.**
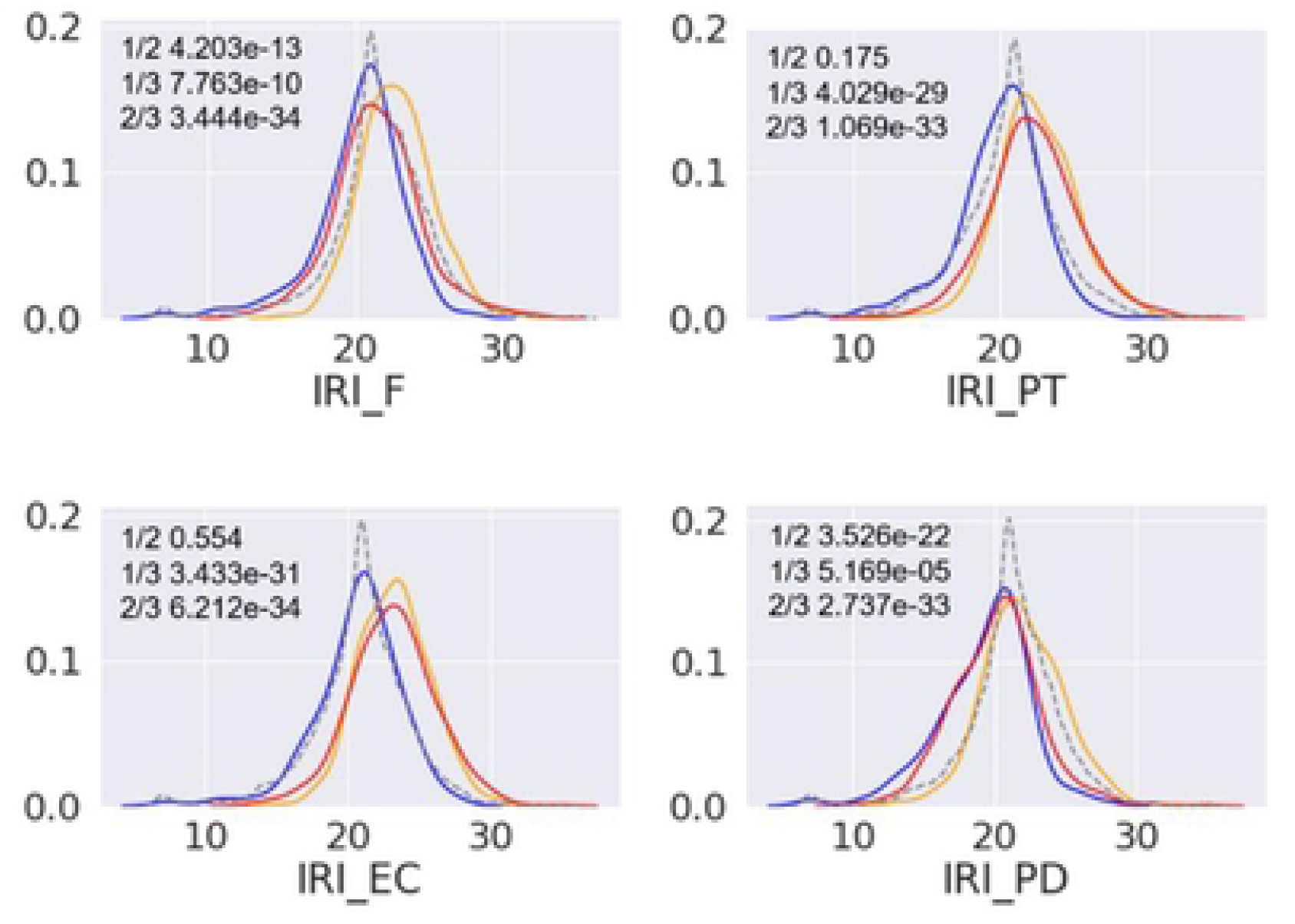
Interpersonal reactivity index for each group. For IRI_F and IRI_PD, Group 2 had significantly higher mean values than Groups 1 and 3 (IRI_F: Group 2 vs. 1, f=-7.350, p=4.203e-13; Group 2 vs. 3, f=12.852, p=3.444e-34; IRI_PD: Group 2 vs. 1, f=-9.939, p=3.526e-22; Group 2 vs. 3, f=12.625, p=2.737e-33). For IRI_PT and IRI_EC, Group 3 had significantly lower mean values than Groups 1 and 2 (IRI_PT: Group 3 vs. 1, f=11.649, p=4.029e-29; Group 3 vs. 2, f=12.712, p=1.069e-33; IRI_EC: Group 3 vs. 1, f=12.127, p=3.433e-31; Group 3 vs. 2, f=12.791, p=6.212e-34). IRI_F: Imagination, IRI_PT: Perspective Taking, IRI_EC: Empathic Concern, and IRI_PD: Personal Distress.

## Discussion

The AQ is a valuable tool for quickly quantifying a particular individual on the ASD continuum. However, there has not been sufficient consideration into different subgroups in the population with high scores. In this study, using a large set of personality traits and attributes other than happiness and the Big5, we conducted a clustering analysis of a large population with high AQ scores not limited to clinical patients. As a result, we identified three subgroups.

Group 1 has high AQ_S, AQ_A, and AQ_C. People in this group tend to be obsessive about one thing, unsociable and not good at communication. Many are paranoid and distrustful, short-tempered, and have low resistance to violence. Depressive and anxiety tendencies are also high, and happiness and self-esteem are lower than those of the overall general population. They also have a tendency toward OCD and schizophrenia: they are highly afraid of failure. Characteristics of Group 1 such as low self-esteem [33] and high hostility [34] along with high SPQ scores [35] are reminiscent of BPD. BPD is estimated to be present in about 1-2% of the general population and more common in females than in males. BPD is often associated with impulsivity, depression, anxiety disorders, paranoid personality disorder, and difficulties in interpersonal relationships [36]. The DSM-IV criterion for BPD is to satisfy five out of nine items, including inappropriately intense anger or difficulty at controlling anger, chronic feelings of emptiness, and self-injurious behavior (which was difficult to confirm in the present study). It is also reported that BPD is characterized by unstable personality traits [37], suggesting that a longitudinal study may be necessary.

Group 2 has high AQ_S and AQ_D, is unsociable, and sensitive to details and anxiety is slightly higher than the overall average. This group is eager to try what they want but afraid of failure. The unique feature of this group is that many of them score high on BIS, IRI and the Big5 (particularly for agreeableness and openness).

Group 3, like the other two groups, has higher AQ_S, SQ_A, AQ_C, and AQ_I scores than the general population, but its AQ_A and AQ_D scores are lower and AQ_I score higher than the other two groups. Overall, most of the scores in this group are comparable with the general population, but among the three groups, Group 3 has the lowest Big5_N, suggesting that people in this group are emotionally stable. The scores of emotionally related items, such as depression and anxiety, are also low. Group 3 has high interpersonal factors of schizophrenia related to social interactions and AQ in general, but its members are emotionally stable and resemble typical Asperger’s syndrome.

Schwartzman et al. [13] conducted a clustering analysis of people with high ASD scores measured by the RAADS-R based on the Big5 and their subdivisions. They identified four clusters. Cluster 1 and Cluster 2 had Big5 traits patterns typical of social phobia [11] and secondary psychopaths and BPD [12], respectively. Cluster 3 had low agreeableness, extraversion, and average levels of neuroticism and conscientiousness but high well-being, and Cluster 4 had the highest extraversion and agreeableness and high well-being.

Group 1 in the present study has high depressive tendencies and anxiety and low happiness. It also had high scores for OCD, schizophrenia, impulsivity, and other strong characteristics of BPD, but not for secondary psychopathy. These data suggest that Group 1 in the present study and Cluster 2 in Schwartzman et al. [13] generally correspond to each other, implying a deeper association with BPD. On the other hand, Group 2 in the present study has high BIS and IRI. High BIS suggests features of depression and social phobia [38, 39], which are closely related to Cluster 1. In addition, social phobia is characterized by high IRI [40–42], while strong connection with IRI has not been reported for depression. Also considering that BDI for Group 2 was not high, we think that Group 2 is more related to social phobia than depression. The present study utilized a large set of personality traits and attributes beyond the Big5, including BIS, which enabled us to conduct a detailed analysis of the AQ based on multiple indices of personality traits and attributes.

By contrast, none of the clusters in Schwartzman et al. [13] matched Group 3, which represents a typical autistic person with a high AQ with other personality scores comparable to the general population. A potential reason for the gap is that only the Big5 and a limited number of indices, such as happiness and life satisfaction, were considered by Schwartzman et al. [13]. In addition, this previous study collected subjects through a website focusing on autism, which may also account for the difference. Related to this, the fact that the gender ratio in the high AQ group of the present study did not match the general 4:1 ratio of males to females for autism may arise from the gender bias in Internet users.

It is possible that the characteristics of personality disorders defined in the DSM5 (i.e., Cluster A-C) provide useful insights into our clustering results. Group 1, which has strong characteristics of BPD, is like Cluster B, which includes BPD, Narcissistic Personality Disorder, Antisocial Personality Disorder, and Acting Personality Disorder, and is characterized by acting, emotional, and volatile characteristics. Among these disorders, people with BPD are considered to have unstable emotions and interpersonal relationships and difficulty controlling impulses. Although we showed high OCD and SPQ scores for Group 1, more study is needed for impulsivity in Group 1. Group 2, which shows characteristics of high suspicion and high anxiety, is like Cluster C, which has anxious and introverted characteristics. Cluster C includes Dependent, Obsessive Compulsive, and Avoidant Personality Disorders. Group 2 also has similar characteristics for avoidant personality disorder, which is characterized by an extreme fear of failure and of being rejected by others. Finally, Group 3, which has high autistic tendencies but average-level scores for traits other than interpersonal relationships, mostly fits into Cluster A. This category includes personality disorders such as paranoid (delusional) personality disorder, schizoid personality disorder, and schizotypal personality disorder. Among these, Group 3 was closest to schizoid personality disorder, which is characterized by a lack of interest in others, a preference for being alone, and a lack of concern for the evaluation of oneself by those nearby.

There are several limitations in the present study. First, although inattentive and hyperactive behaviors are common in children with ASD, children with ADHD also show similar social difficulties with their peers [43]. Before the development of the DSM-5, it was not possible for a person with a diagnosis of ASD to be simultaneously diagnosed with ADHD. However, studies after DSM-5 have shown that ADHD is the most common comorbidity among children with ASD, with a probability of 40%-70%. In addition, ADHD and ASD have many copy number variants (CNV) and chromosomal abnormalities in common, indicating genetic overlap between them [44]. Therefore, it is necessary to include ADHD measures in large-scale studies of ASD in the future.

Second, although our results revealed heterogeneity in the high AQ samples, this does not necessarily mean that people with ASD comprise three subgroups. It is possible that AQ cannot separate autism from other psychiatric disorders. Even so, we believe our results will contribute to the better understanding of AQ.

Third, although we confirmed that the number of clusters was three by using two different metrics for clustering, the cluster size may differ depending on the methods and data, as discussed in a previous study on personality types based on large-scale data [16], which reported that a simple application of clustering yields a spurious number of personality types. Therefore, we took a minimalist approach to increase the cluster size gradually. That is, when we first employed the K-means clustering method instead of the Ward method, we found two clusters: one with high scores for mental illness and the other with low scores. This result is consistent with a previous clustering study [13] of high autistic tendency groups based on the Big5, which reported one group with characteristics like mental disorders and the other with only high autistic tendencies. Since we were mainly interested in the structures in the high-score group, we decided to adopt the Ward method in the present study.

Fourth, the personality test data of this study was collected from adult Japanese speakers between the ages of 19 and 89, and all participants satisfied the condition to have cognitive ability to complete the surveys through internet access. Different clusters may emerge if recruiting non-Japanese or children-only participants.

In summary, this study conducted a clustering analysis of a large number of high AQ subjects based on a large set of personality traits and attributes and identified three subgroups: Group 1, which resembles borderline personality disorder; Group 2, which resembles social phobia; and Group 3, which has high AQ scores but other personality scores that resemble the general population. These findings lead to several important questions for future study. The first is to address whether the three subgroups are also seen in the clinical population of ASD and how they show differences in real social behaviors. A previous study based on the Big5 scores from 64 Asperger’s syndrome patients reported larger variances in the patients than in normal controls, potentially indicating the existence of subgroups [45]. The second is to address how AQ scores and other personality measures are distinct and overlapping [10]. We believe these new avenues based on a data-driven approach like ours will help clarify AQ and ASD.

## Materials and Methods

### Participants

Subjects older than 18 years old were recruited through an online survey over a four-year period from 2016 to 2019. A total of 18926 subjects (10416 males and 8510 females) whose ages ranged from 18∼89 years old were collected for the analysis, excluding duplicate subjects who took the test multiple years. The high AQ group consisted of 1364 subjects (819 males and 545 females) whose ages ranged from 19 and 83 years old. The mean age was 39.0 years old, with a standard deviation of 14.7 years. Because all personality questionnaires were written in formal Japanese, proficiency in the Japanese language was required to complete the survey. Therefore, all participants were likely to be Japanese, although some may have been foreigners fluent in the language. The online survey and analysis methods were reviewed and approved by the Ethics Committee of National Institute of Information and Communications Technology. The methods were in accordance with the code of ethics and conduct of the Japanese Psychological Association. Subjects were briefed about the experiment, and informed consent was obtained from all. The research questions and analyses were not preregistered.

### Online survey

Data collection was conducted through an online survey in Japanese as follows. A research company recruited the subjects and issued a unique URL to each of them. Subjects accessed the personality trait questionnaires on the server at our institute through the received URL. At the end of the test, we sent a “linkage ID” of the subject with the completion status of the questionnaire to the research company. Based on this ID, the research company gave the subjects reward points (equivalent to 300 yen).

The contents and abbreviations of the personality trait questionnaires are explained in Table S1. Specifically, the mental health scores included schizophrenia [SPQ_C, SPQ_I, SPQ_D], delusion [PDI_DE, PDI_DI, PDI_FR, PDI_CO], psychopathy [PSPS_P, PSPS_S], OCD [OCIR_WA, OCIR_OB, OCIR_ST, OCIR_CH OR, OCIR_CH, OCIR_CH], depression [BDI, SDS], anxiety [STAI_S, STAI_T], and stress [PSS]. (The abbreviations in square brackets indicate subscales.) Behavioral economics scores included socioeconomic measures [SES] and social value orientation [SVO_P, SVO_I, SVO_C]. Money-related risk aversion and time discounting [RA, TIM] were also examined. Empathy-systematization scores included scales for empathy [IRI_F, IRI_PT, IRI_EC, IRI_PD] and ASD [AQ_S, AQ_D, AQ_A, AQ_C, AQ_I]. The inhibition/activation score included scales for the Behavioral Inhibition System (BIS) and the Behavioral Activation System (BAS) [BAS_D, BAS_R, BAS_S]. All questionnaires were self-reported as described in S1 Table.

### The Autism-Spectrum Quotient (AQ)

We used a Japanese version of the AQ [46], which comprises 10 items for each of five major domains (social skills, attention switching, attention to detail, communication, and imagination). The AQ uses a forced-choice self-report format. Answers are rated on a binary scale, and the maximum score is 50 points; a higher score indicates stronger autistic tendency. Because the cutoff point between people with AS/HFA and the general Japanese population was previously identified to be 32 points [46], AQ scores of 33 or higher were used to define the high AQ group for clustering.

### Analysis

#### Lasso regression

Initially, to determine which personality traits relate to the AQ, we ran a Lasso regression analysis of scikit-learn in Python [47] to predict individual AQ scores from a total of 56 personality traits (sub-) scores (excluding the AQ) and attributes. In the Lasso regression analysis, we used the total AQ score to characterize the high AQ group beyond the Japanese AQ cutoff value (32). In the subsequent analysis to characterize each high AQ subgroup, we examined subscales rather than total scores to examine differences in subscales among subgroups. Data including a missing value was excluded from the analysis.

#### Clustering

We clustered the high AQ group of 1364 people (7.2% of total respondents) using the Ward method (using hclust in R) with two different metrics: the Euclidean distance and the Manhattan distance. We used 35 personality traits whose Lasso coefficient was larger than 1/20 of the largest absolute value of weight in the Lasso regression analysis. The optimal number of clusters was determined to be 3 by the NbClust method [48] implemented on R for both the Euclidean and the Manhattan distances. The cutree function in R was used to create a tree diagram. We also tested the K-means algorithm for clustering.

#### Score distributions for each group

We drew a kernel distribution estimate plot and examined the distribution of all personality trait scores for each group using kdeplot of seaborn [49] in Python.

## Data Availability

All personality trait scores used in this study will be available from the OPEN ICPSR database.

## Acknowledgements

We would like to thank Satoshi Tada and Dr. Tomoki Haji for technical assistance, and Peter Karagiannis for editing an early version of the manuscript.

## Supporting information

**S1 Fig. Three clusters from the Manhattan distance.**

**S1 Table. The contents, abbreviations and references of the personality trait questionnaires.**

**S2 Table. Oneway ANOVA result.**

## Data and code availability

The data and code that support the findings of this study are available on request to the corresponding author.

